# Characterizing Potential Conflicts of Interest Among UpToDate and DynaMed Content Contributors

**DOI:** 10.1101/2021.10.12.21264909

**Authors:** SooYoung H VanDeMark, Mia R Woloszyn, Laura A Christman, Michael Gatusky, Warren S Lam, Stephanie S Tilberry, Brian J Piper

## Abstract

**BACKGROUND:** Financial conflicts of interest among physicians have the potential to negatively impact patient care. Physicians contribute content to two popular, evidence-based websites, UpToDate and DynaMed; while other physicians use these websites to influence their clinical decision making. Each website maintains a conflict-of-interest policy, and contributors are required to self-report a disclosure status. This research investigated the occurrence for potential conflicts of interest among the self-reported statuses of UpToDate and DynaMed content contributors.

**METHODS:** An initial list of contributors for each website was compiled using the Centers for Disease Control and Prevention’s 2017 Leading Causes of Death. The top 50 causes were used to determine a relevant article with clinical implications from each database. All named authors and editors of those articles comprised our list of investigated contributors. Contributor disclosure status was then compared with public records of financial remuneration as reported in the Open Payments database maintained by the Centers for Medicare and Medicaid Services and ProPublica’s Dollar for Docs website from 2013 to 2018. Descriptive analysis and Fisher’s exact tests were performed on the data.

**RESULTS:** Of 76 UpToDate contributors, 57.9% reported nothing to disclose but had a record of receiving a financial payment on Open Payments, which was found to be statistically significant (p = 0.0002). Of DynaMed’s 42 contributors who reported nothing to disclose, 83.3% had an entry on Open Payments. However, this was not statistically significant. The sum total of industry payments between 2013-2018 made to UpToDate contributors was $68.1 million. The top ten UpToDate contributors who received the most financial remuneration earned approximately $56.1 million (82.4% of all UpToDate renumeration), were all male, and only one had a nothing-to-disclose status. The sum total of compensation reported for the discordant UpToDate contributors between 2013-2018 was approximately $4.81 million (or 7.07% of the total monies reported to UpToDate contributors.) In that same time frame, DynaMed contributors received a sum total of $9.58 million from industry, while the top ten DynaMed contributors earned $8.88 million (or 92.8%) of that. The top ten DynaMed contributors were 80% male and 20% female, and six individuals reported nothing to disclose, yet had an Open Payments entry. The sum total of money reported for all discordant DynaMed contributors between 2013-2018 was approximately $2.79 million (or 29.2% of the total monies reported to DynaMed contributors).

**CONCLUSIONS:** While this research does not ascertain that a conflict of interest or anything untoward, it does provide evidence that there was a significant difference between having an Open Payment entry among those who did versus those who did not disclose a conflict of interest. Websites such as UpToDate and DynaMed should consider implementing a more stringent conflict of interest policy and employ an unbiased team to verify self-reported disclosure statuses among its content contributors. Similarly, physicians who use such informational websites to inform their clinical decision making should look beyond a contributor’s self-reported disclosure status and verify relevant financial remuneration from the healthcare industry via Open Payments or Dollars for Docs.

## Introduction

Per the Institute of Medicine’s Committee on Conflict of Interest in Medical Research, Education, and Practice, a conflict of interest (COI) is defined as a set of “circumstances that create a risk that professional judgments or actions regarding a primary interest will be unduly influenced by a secondary interest” (1). This investigation is focused on investigating potential COIs that could occur when a physician writes or edits medical content (the primary interest), yet is influenced by financial gain (the secondary interest) in the form of payments made by health care manufacturers to the author. Undisclosed COIs may not only affect the credibility of a health resource, but also other clinicians’ ability to provide quality care.

Prior quantitative bioethical research has found potential COIs among physician authors of biomedical textbooks (2), pharmacology textbooks (3), psychiatry’s Diagnostic and Statistical Manual of Mental Disorders-5 (4), as well as within clinical guidelines (5). However, there is limited research on the potential COIs among authors and editors of online clinical resources, such as UpToDate and DynaMed. UpToDate and DynaMed are two online, subscription-based products used by physicians to assist in clinical decision making. Both UpToDate and DynaMed promote their websites as an evidence-based resource for physicians to improve patient health outcomes at the point-of-care (6,7). The content on these websites is written, edited, and overseen by various health care professionals. UpToDate and DynaMed each maintain a publicly available COI policy (8,9).

In an effort to improve transparency and as part of the Physician Payments Sunshine Act, the Centers for Medicare and Medicaid Services (CMS) discloses financial payments from “drug, device, biological, and medical supply” manufacturers to U.S.-based physicians through its online and publicly-accessible database, Open Payments (OP) (10). Another online, publicly-accessible database is Dollars for Docs (DFD), which is maintained by the nonprofit, investigative journalism organization, ProPublica. DFD also allows users to look-up payments made from relevant manufacturers to U.S.-based physicians as reported to CMS (11). OP and DFD only report payments in amounts greater than $10.

As mentioned, there has been limited research on potential COIs among authors of evidence-based, point-of-care websites. Researchers reviewed authors (n=31) of the online medical resource, Medscape, for potential COIs and found 19 discordant authors (61.3%) who self-reported a nothing to disclose status, but had an OP entry (12). Additionally, this investigation (12) found a strong correlation between the reported payment amounts on OP and DFD, consistent with prior research (3). A small report focused on UpToDate and DynaMed authors and editors, reviewed six articles on each site and found no COIs among DynaMed contributors and “numerous” potential COIs among UpToDate contributors (13).

We investigated potential COIs among content contributors for DynaMed and UpToDate by cross-checking their self-reported disclosure statuses with financial records available from OP and DFD. Additionally, a descriptive analysis of UpToDate and DynaMed content contributors’ disclosure status, financial compensation, and gender was performed, with further evaluation of each website’s top ten earners.

## Methods

### Procedures

UpToDate and Dynamed were selected based on their favorable ratings in empirical evaluations of breadth of coverage, timeliness of updating, use of evidence-based methodology, and utility (14-16) and also because our library subscribed to them. Using the Center for Disease Control’s Top 50 Causes of Mortality (17), each cause of death was searched for on UpToDate’s and DynaMed’s website. The research team, with individuals assigned to specific diseases, selected comparable articles from the first page of search results on each site. Only 42 causes of death were used for data collection, resulting in a total of 84 articles reviewed. Diseases were excluded from data collection, if they provided no relevant search result on UpToDate or DynaMed (e.g., “operations of war and their sequelae.”)

Content contributors were defined as the individuals listed on a given UpToDate or DynaMed article page—specific titles for contributors depend on the database, but include: Author, Deputy Editor, Section Editor, Recommendations Editor, and American College of Physicians (ACP) Reviewer. Each article’s listed content contributors, regardless of title, composed our initial list of physician contributors. Contributors who were of unknown or international origin were removed from this study, as OP and DFD only report on U.S.-based physicians. A contributor states nothing to disclose or discloses the companies from which they have received payment under each article on both websites. This status was collected. All articles and contributor names were compiled between 11/30/2020 and 12/07/2020.

Each unique contributor (n=179, 28.5% female) was then searched for in OP and DFD. If an entry was found, the financial information for 2013-2018 was recorded in accordance with each website’s categorization method. For example, money reported to OP was categorized as General Payments, Research, Associated Research and Owner/Investment. Contributor location and gender was verified or cross-checked and documented using the National Provider Identifier registry website (15).

The research team performed a randomized check on 11.2% of the data collected to ensure accuracy. No individuals were found to have contributed content to both UpToDate and DynaMed. Although some contributors were listed on multiple articles, no variability in disclosure status was found between articles. After data collection and prior to analysis, contributors were given a unique ID to avoid potential bias from the research team. Contributors were identified as discordant if they reported nothing to disclose, but were found to have an OP or DFD entry. Contributors were identified as concordant if they made a disclosure, and had an OP or DFD entry.

### Statistical Analysis

Two-sided Fischer exact tests were performed, due to the study’s small sample sizes, using Prism 9.1.0 (16). This study was deemed exempt from review by the Geisinger IRB.

## Results

Of the 179 U.S-based physician content contributors, associated with the 84 articles, 128 were from UpToDate and 51 were from DynaMed (Figure 1). The sum total of reported financial compensation to OP for the UpToDate contributors, between the years 2013-2018, was $68,085,233; of which, the top ten earners accounted for $56,083,923 or 82.4%. The sum total reported to OP for the DynaMed contributors, within that same time frame, was $9,576,109; of which, the top ten earners accounted for $8,882,249 or 92.8%.

**Figure 1.**
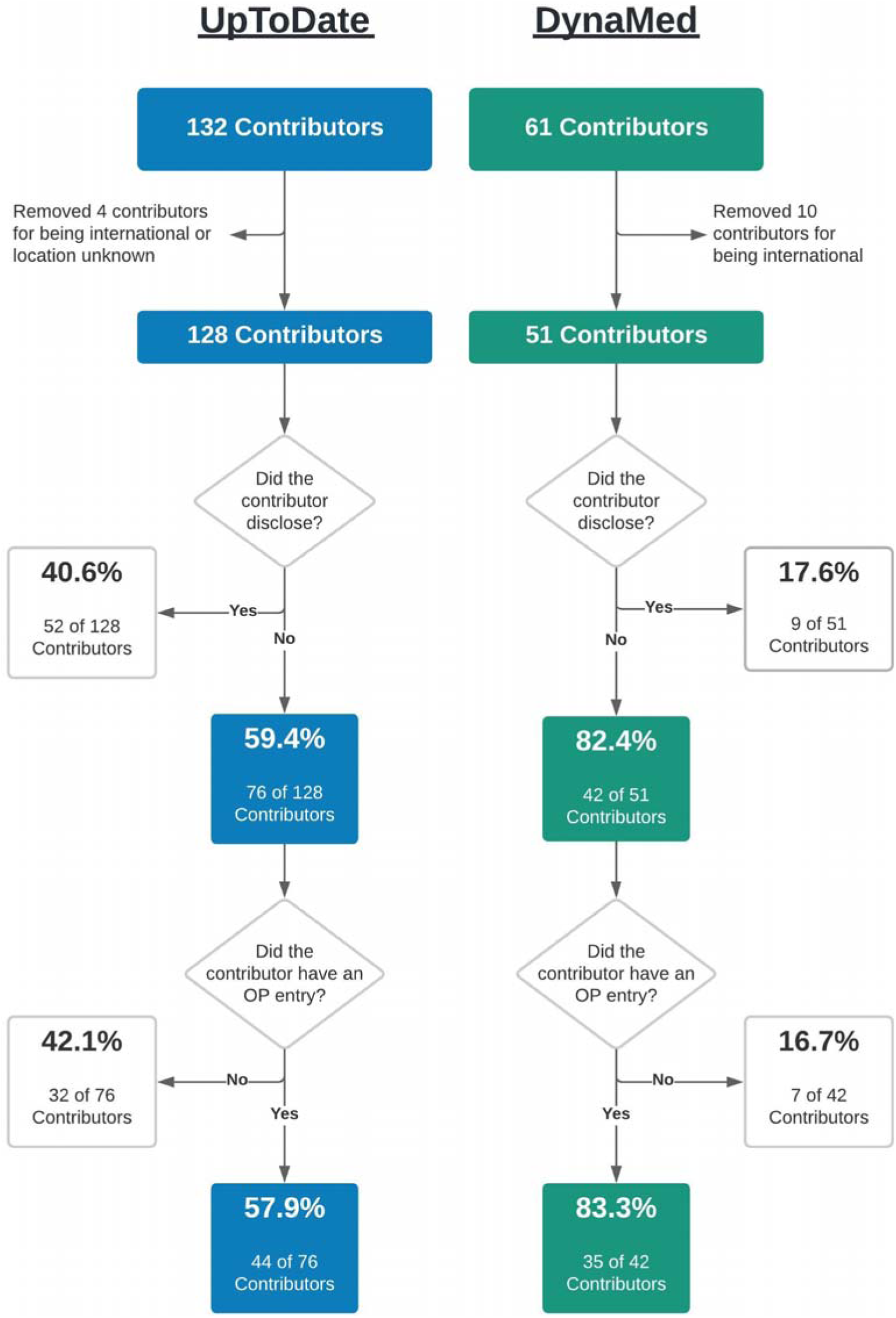
Flow chart of UpToDate (left) and DynaMed (right) content contributors, showing disclosure status and Open Payment (OP) entry status.

The majority of UpToDate (59.4%) and DynaMed (82.4%) contributors did not disclose any COIs related to their article topic (Figure 1). However, of those UpToDate contributors who did not disclose a COI, 57.9% had an OP entry. This discordance— self-reporting nothing to disclose, yet having an OP entry—among UpToDate contributors was statistically significant (p = 0.0002). The 44 discordant UpToDate contributors accounted for $4,811,760 or 7.07% of UpToDate’s sum total as reported by OP. Although 83.3% of DynaMed’s nothing-to-disclose contributors had an OP entry, it was found to be not statistically significant. The 35 discordant DynaMed contributors accounted for $2,793,708 or 29.2% of DynaMed’s sum total as reported by OP.

Six content contributors for UpToDate were found to be neither discordant nor concordant. They had made a disclosure, but no OP nor DFD record was found. Each of the nine DynaMed content contributors who disclosed were found to be concordant.

All of the top ten earners within the UpToDate content contributors were male, of whom only one contributor was found discordant (Figure 2). There were six discordant contributors among the top ten earners of DynaMed contributors, of whom eight were male and two were female (Figure 2). Figure 2 also shows a breakdown of the reported compensation (in millions) to the top ten earners of each website respectively in the four OP categories, with Associated Research, which is defined as “funding for a research project or study where the physician is named as the principal investigator”^10^ dominating the payments. Further investigation of the seven discordant top ten earners found one contributor who received payment(s) from a manufacturer for drug(s) and/or medical device(s) that was specifically mentioned by brand name in the article to which the contributor was assigned. The financial renumeration to this contributor was $4,695.

**Figure 2.**
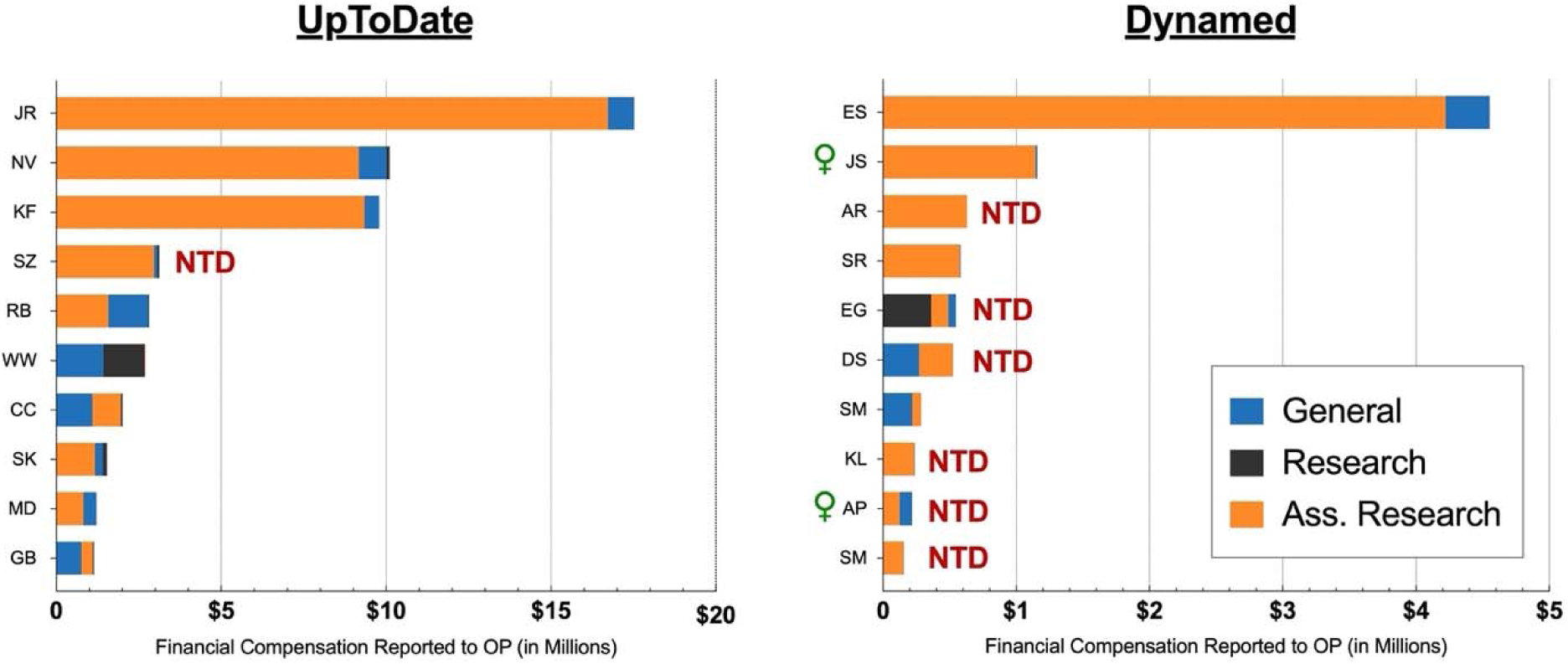
Financial remuneration (in millions) by category as reported to Open Payments (OP) for the top ten earners among UpToDate (left) and DynaMed (right) content contributors. NTD = Nothing to disclose. Female symbol designates the two female contributors.

Among all UpToDate content contributors, 94 (73.4%) were male and 34 (26.6%) were female. A Fischer’s exact test found a statistically significant difference in that male UpToDate contributors (47.9%) were more likely to report a disclosure than females (20.6%, p = 0.0076). Among the DynaMed contributors, 34 (66.7%) were male and 17 (33.3%) were female. There was no statistically significant difference in how the genders disclosed among DynaMed contributors. Another Fischer’s exact test found no statistically significant difference in discordance based on the contributor’s gender for either website.

## Discussion

This study identified appreciable financial COIs among point-of-care contributors with potential room for improvement in self-reported disclosures. In contrast to prior research (12), the present investigation found potential COIs among contributors for both online resources, UpToDate and DynaMed. This is likely due to a difference in methodologies. Our findings of discordance in UpToDate and DynaMed (57.9% and 83.3% respectively) are similar to a prior finding of 61.3% discordance among Medscape’s contributors (n=31) (11). Such high discordance rates suggest a need for further research to fully illuminate the issue, as well as follow-up remediation by these point of care online clinical information resources.

Based on the high percentage of discordance found among content contributors for both UpToDate and DynaMed, and the large sums of remuneration among those discordant (over $7 million) in this study, there is a strong likelihood for there to be potential COIs among physician content contributors who self-report. Our own investigation into a subgroup of our sample contributors found evidence suggestive potential COI. However, it is important to note that the objective of this research was not to ascertain any specific contributor’s COI.

Our gender analysis revealed an interesting difference in disclosure status between males and females. However, there are multiple ways to interpret this—perhaps, male physicians disclose more because they are more often the beneficiaries of industry dollars and in a position requiring disclosure; or they are more ethical than female physicians; or perhaps, the difference was due to the underrepresentation of female physician content contributors, which has been confirmed in prior studies regardless of the source being a printed text or an online website (2, 3, 12).

One argument against the findings of this research may be that as physicians contribute content to reputable online point-of-care websites, such as UpToDate and DynaMed, they might then be hired by the healthcare industry for their expertise and financially compensated, all within the same calendar year. In this plausible example, no direct COI has occurred. Importantly, OP and DFD provide specific dates for the payments reported to a physician but no such timeline is provided by UpToDate or DynaMed on when the content was initially published. Each site does provide a last revised date but not when the contributor declared their disclosure status. In order to work within these confines, the research team opted to focus on disclosure status and existence of an OP/DFD entry rather than timeline.

Additional limitations of this study include that the sample size of contributors, although focusing on diseases and conditions of substantial importance, was only moderate (N = 179). The objective was not to ascertain the number of contributors for all the content on either UpToDate or DynaMed or the veracity of all disclosures but only a carefully selected subset of the top fifty causes of death in the US. These findings are limited to two widely-used point of care databases (16) and may not generalize to other databases (15) or non-English databases. Similarly, our data is only as accurate as the financial reporting provided by OP and DFD. Given that a small subset of contributors (3.35%) were neither discordant nor concordant, this may be of concern worthy of further empirical attention.

## Conclusion

Our recommendations for evidence-based, point-of-care websites are two-pronged. First, the disclosure information provided for each contributor could be made more robust by providing a “verified” date or timeframe for a nothing-to-disclose status by hyperlinking to the OP and DFD pages for those who have entries and by displaying a monetary range of financial compensation (e.g., $5,000 - $10,000) for those who have OP and/or DFD entries. Such changes would offer transparency to the website user who consumes the information. Second, current COI policy should be reviewed and updated annually, and a verified “no-COI” editorial team should be established to crosscheck physician contributors at random against OP and DFD. Such a policy might result in content contributors erring on the side of caution and disclosing more openly and completely. Although the dearth of female authors was consistent with prior studies, further study should examine whether this scarcity impacts how or what content is presented in these widely utilized point of care databases.

Research in the area of medical authorship and COIs needs to continue, with a particular emphasis placed on online medical resources. Physicians, other health care providers, and by extension their patients, should have maximal confidence knowing that the evidence-based medical information they receive is free from outside influences.

## Data Availability

Raw data is available at: https://projects.propublica.org/docdollars/

## Acknowledgements

The research team would like to thank Kaitlyn E Sternat and Amalie Kropp Lopez for their contributions to this project. BJP is supported by the Health Resources Services Administration (D34HP31025).

